# Lipoprotein and metabolite associations to breast cancer risk in the HUNT2 study

**DOI:** 10.1101/2021.10.08.21264729

**Authors:** Julia Debik, Hartmut Schaefer, Trygve Andreassen, Feng Wang, Fang Fang, Claire Cannet, Manfred Spraul, Tone F. Bathen, Guro F. Giskeødegård

**Affiliations:** Department of Circulation and Medical Imaging, Norwegian University of Science and Technology, Trondheim, Norway; Bruker BioSpin AIC Division, Ettlingen, Germany; Clinic of Surgery, St. Olavs Hospital, Trondheim University Hospital, Trondheim, Norway; Department of Radiology and Nuclear Medicine, St. Olavs Hospital, Trondheim University Hospital, Trondheim, Norway; K.G. Jebsen Center for Genetic Epidemiology, Department of Public Health and Nursing, Norwegian University of Science and Technology, Trondheim, Norway

**Keywords:** Breast cancer, Prediagnostic serum, NMR spectroscopy, Lipoproteins

## Abstract

**Background:** The aim of this study was to investigate if serum lipoprotein and metabolic profiles of healthy women can predict the risk of developing breast cancer in the future, and to gain a better understanding of the etiology of the disease.

**Methods:** From a cohort of 70 000 participants within the Trøndelag Health Study (HUNT study), we identified 1199 women who developed breast cancer within a 22 year follow-up period. Through a nested case-control study design, future breast cancer patients and matching controls (n = 2398) were analysed. Using nuclear magnetic resonance (NMR) spectroscopy, 28 metabolites and 112 lipoprotein subfractions were quantified from prediagnostic serum samples. Logistic regression was used to test metabolites and lipoprotein subfractions for associations with breast cancer risk and partial least-squares discriminant analysis (PLS-DA) models were built to predict future disease.

**Results:** Among premenopausal women (554 cases) 14 lipoprotein subfractions were associated with long-term breast cancer risk. In specific, different subfractions of VLDL particles (in particular VLDL-2, VLDL-3 and VLDL-4) were inversely associated with breast cancer. For total VLDL: apolipoprotein B, cholesterol, free cholesterol and phospholipids were inversely associated with premenopausal breast cancer risk, and in addition total and HDL-4 triglycerides. No significant association was found in postmenopausal women.

**Conclusions:** We identified several associations between lipoprotein subfractions and long-term risk of breast cancer in premenopausal women. Inverse associations between several VLDL subfractions and breast cancer risk were found, revealing an altered metabolism in the endogenous lipid pathway many years prior to a breast cancer diagnosis.

## Introduction

Breast cancer is the most common cancer diagnosis among women, and the incidence rate is following an increasing trend [1]. Although the overall breast cancer survival rates have increased bringing the 5-year survival up to 90.7% in Norway [2], the prognosis greatly depends on the stage of the disease at diagnosis, in addition to treatment efficacy. A better understanding of the etiology of the disease and the biological mechanisms leading to disease could reveal methods for disease prevention and early detection.

Cancer cells have a reprogrammed metabolism for conversion of nutrients to biomass while maintaining a high energy production. This phenomenon is increasingly recognized as a source for biomarkers for early detection. The serum metabolome is affected by the preceding levels of the omics cascade and external factors, providing a detailed snapshot of the current state of the organism [3-5]. Significant changes in the metabolism of breast cancer patients have been described, both in tumor tissue and biofluids [6-10]. Furthermore, subtle differences in metabolic composition of prediagnostic serum samples have been associated with breast cancer risk [11-18].

Lipids, playing a critical role in cell signaling and membrane formation, have altered levels in many types of cancer [19, 20]. However, the mechanisms governing dysregulated lipid metabolism in cancer development are not fully understood. The two main forms of circulating lipids in the body are triglycerides and cholesterol, which are transported through the bloodstream in lipoproteins. Lipoproteins are particles with an inner core, mainly composed of triglycerides and cholesteryl esters, surrounded by a hydrophilic membrane consisting of free cholesterol, phospholipids and apolipoproteins [21]. There are five main fractions of circulating lipoproteins, ranging from very-low (VLDL) to high-density (HDL) lipoproteins, each with its own characteristic protein and lipid composition. Traditional lipoprotein measurements, however, do not capture the delicate density range within the main fractions. Detailed characterization of the different lipoprotein subfractions and their content, possible through nuclear magnetic resonance (NMR) spectroscopy, may give important biological information on early breast cancer development.

In this study we aimed to identify associations between lipoprotein subfractions and circulating metabolites in prediagnostic serum samples and breast cancer risk, and to gain insight into the etiology of the disease. Our study aims were accomplished by a case-control study nested in the Trøndelag Health Study (the HUNT study), with samples taken up to 22 years before breast cancer diagnosis.

## Materials and methods

### Sample Collection and Experimental Design

The Trøndelag Health Study (the HUNT study) is a longitudinal population health study in Norway, including 230 000 participants. It includes a database of questionnaire data, clinical measurements, and biological materials. So far four health surveys have been completed, HUNT1 (1984-86), HUNT2 (1995-97), HUNT3 (2006-08) and HUNT4 (2017-19) [22] in addition to an adolescent part (13 – 19 years) [23]. HUNT2 was the first study to include biological material. By matching data from HUNT2 with the Norwegian Cancer Registry, we have identified all participants of HUNT2 that developed breast cancer between data collection and follow-up in 2019. The mean time from blood collection to a breast cancer diagnosis was 11.7 years (range 0-22 years). For each case, a participant that remained breast cancer free during follow-up was randomly selected as a control, matched for age at inclusion into HUNT2 in intervals of 5 years. Relevant clinical variables were selected from the HUNT2 databank for both the cases and controls, while cancer-specific variables were retrieved from the Norwegian Cancer Registry. All participants have completed a written informed consent form, and the study was approved by the Ethics Committee of Central Norway (REK numbers #1995/8395 and #2017/2231).

### NMR Experiments and metabolite quantification

Serum samples were stored at -80°C until analysis and NMR experiments were carried out at two separate labs, located in Norway (MR Core facility, NTNU) and Germany (Bruker BioSpin GmbH). The study cohort included 1199 cases and 1199 matched controls, and the selection of samples to be analyzed at the two labs was random. The NMR spectrometers at the both labs have been calibrated for use of the same protocol (Supplementary Methods). NMR spectra were recorded using one-dimensional nuclear Overhauser effect spectroscopy (1D-NOESY) and Carr-Pucell-Meiboom-Gill (CPMG) experiments. The absolute metabolite concentrations were obtained for a total of 28 from CPMG spectra with correction for T2 attenuation (Table S1, Supplementary Methods). The coefficients of variation (CVs) of the metabolites were below 15% and below 20% for 23 and 26 of the metabolites, respectively (Table S1). Lipoprotein subfractions were automatically quantified using Bruker IVDr Lipoprotein Subclass Analysis (B.I.LISA^™^) software, from Bruker BioSpin. This software provides the concentrations of lipids [cholesterol (CH), free cholesterol (FC), triglycerides (TG), and phospholipids (PL)] in total serum and in four main lipoprotein classes: very low-, intermediate-, low-, and high-density lipoproteins (VLDL, IDL, LDL, and HDL) and 15 subclasses (VLDL 1-5, LDL 1-6, and HDL 1-4). In addition, serum levels of apolipoproteins (Apo-A1, Apo-A2, and Apo-B) in the lipoproteins, 12 calculated parameters (ratios of LDL-CH/HDL-CH and Apo-B/Apo-A1), and particle numbers of total serum, VLDL, IDL, LDL, and LDL 1-6 are provided, giving a total of 112 lipoprotein subfractions. However, due to the presence of a contamination in the serum samples (neopentyl glycol, most likely originating from the original collection tubes), which interfered with the broad lipid peak at 0.85 ppm on the ^1^H spectra, some of the lipoprotein subfractions were excluded from further analysis (Supplementary Methods, Table S2, Figure S1), mostly from LDL-2 and LDL-4 particles. Calculated parameters and particle numbers were also excluded from the statistical analysis, resulting in 89 lipoprotein subfractions. CVs for the lipoprotein subfractions included were below 15% and 20% for 65 and 85 of the variables, respectively (Table S2).

### Statistical analysis

Baseline characteristics of the cases and controls were described using mean and standard deviation (SD), and statistical significance was assessed by Student t-tests for continuous variables and the Fisher’s exact test for categorical variables.

NMR derived variables were standardized to unit variance prior to statistical analysis. Correlation between the NMR-derived variables was tested using Pearson correlation analysis. Odds ratios (OR) and 95% Wald confidence intervals (CI) were calculated for a one SD increase in the concentration of each variable using logistic regression. P-values were corrected for multiple testing using the Benjamini-Hochberg approach, and significance was considered for P_adj_ ≤ .05. The baseline model was adjusted for age (matching factor) and lab at which the NMR measurement took place, due to the presence of a batch effect between labs. In the adjusted model, the following additional factors were included to correct for confounding factors: number of full-term pregnancies, age at menarche, body mass index (BMI), alcohol consumption (monthly frequency) and smoking status (current smoker or non-smoker). Menopausal age was available for only 483 of the individuals, all remaining individuals aged 51 or higher at participation in HUNT2 were classified as postmenopausal. This cut-off value was chosen at the basis of a large population study in Norway, including more than 300 000 individuals [24]. Clinical variables with less than 10% missing values for the cohort were included in the logistic regression model, where the missing values were imputed with the median values of the full cohort. The use of hormone replacement therapy (HRT) at baseline (systemic, local, previous or never use) was missing for 20.8% of the study cohort, thus this variable was not included in the adjusted model. To evaluate the influence of HRT usage on the studied associations, we concluded stratified analyses excluding women for whom use of HRT was reported or the information was missing. Stratified analyses were also performed for estrogen receptor positive (ER+) and negative (ER-) breast cancers.

Multivariate predictive models were fit using partial-least squares discriminant analysis (PLS-DA) for discriminating between lipoprotein profiles of pre- and postmenopausal women, and between cases and controls in pre- and postmenopausal women separately. The models were validated using double cross-validation, and their significance (P_perm_ ≤ .05) was assessed by permutation testing (1000 permutations). Stratified analysis, based on the number of years between sample collection and breast cancer diagnosis were also performed.

## Results

### Population characteristics

The baseline characteristics of the clinical and lifestyle variables for the participants are summarized in Table 1. Considering traditional breast cancer risk factors, there were significant differences between the groups for age at first pregnancy, the number of full-term pregnancies, height and alcohol intake (p < .05). The controls had their first full-term pregnancy at a younger age compared to the cases, and a higher number of full-term pregnancies. The frequency of alcohol intake was significantly higher for the cases. Women who developed breast cancer were also significantly taller than controls, while there was no difference in BMI and waist-hip ratio (WHR). Cases had more often diabetes and use of systemic HRT, however the difference between the cases and controls did not reach statistical significance.

**Table 1.**
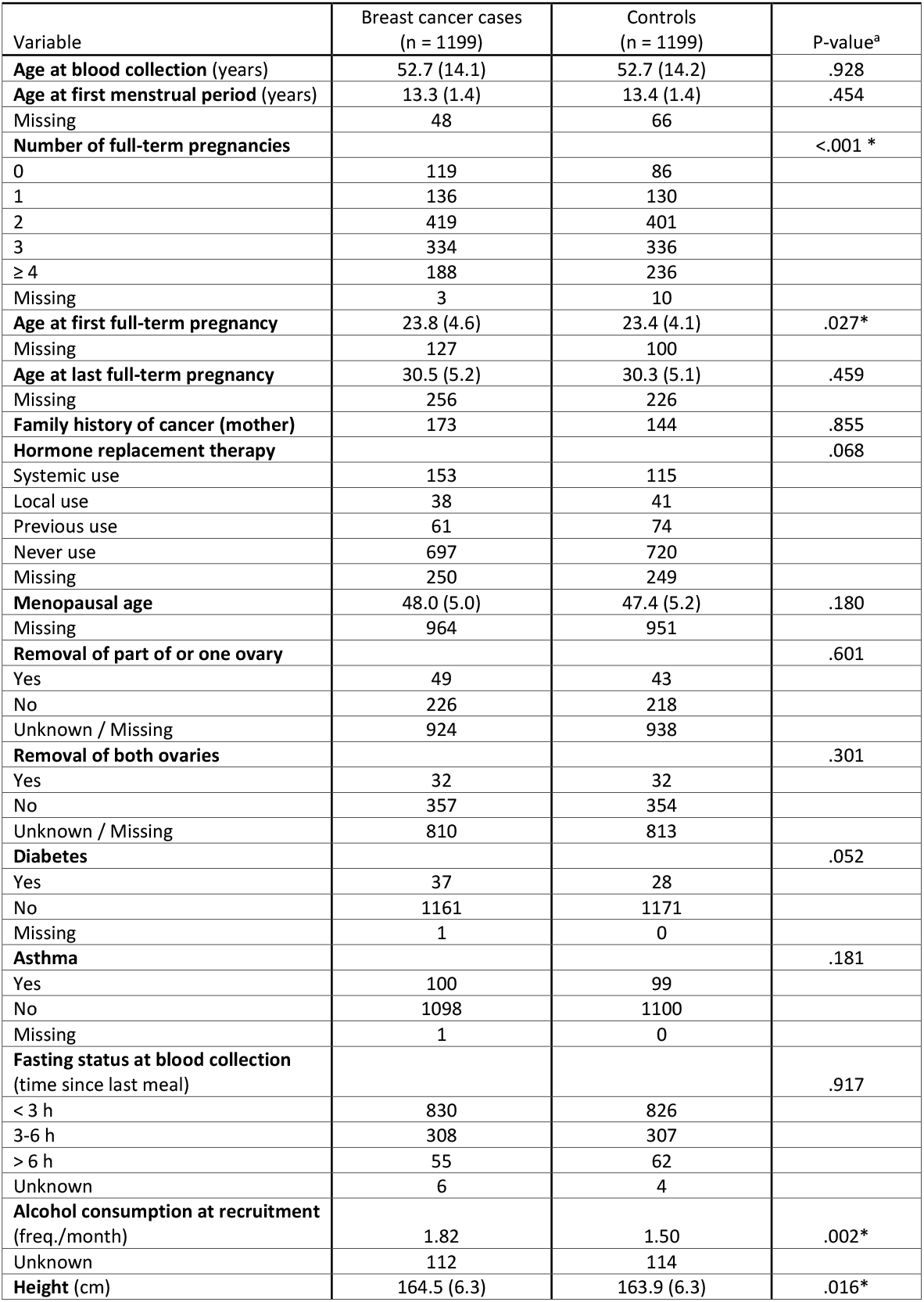
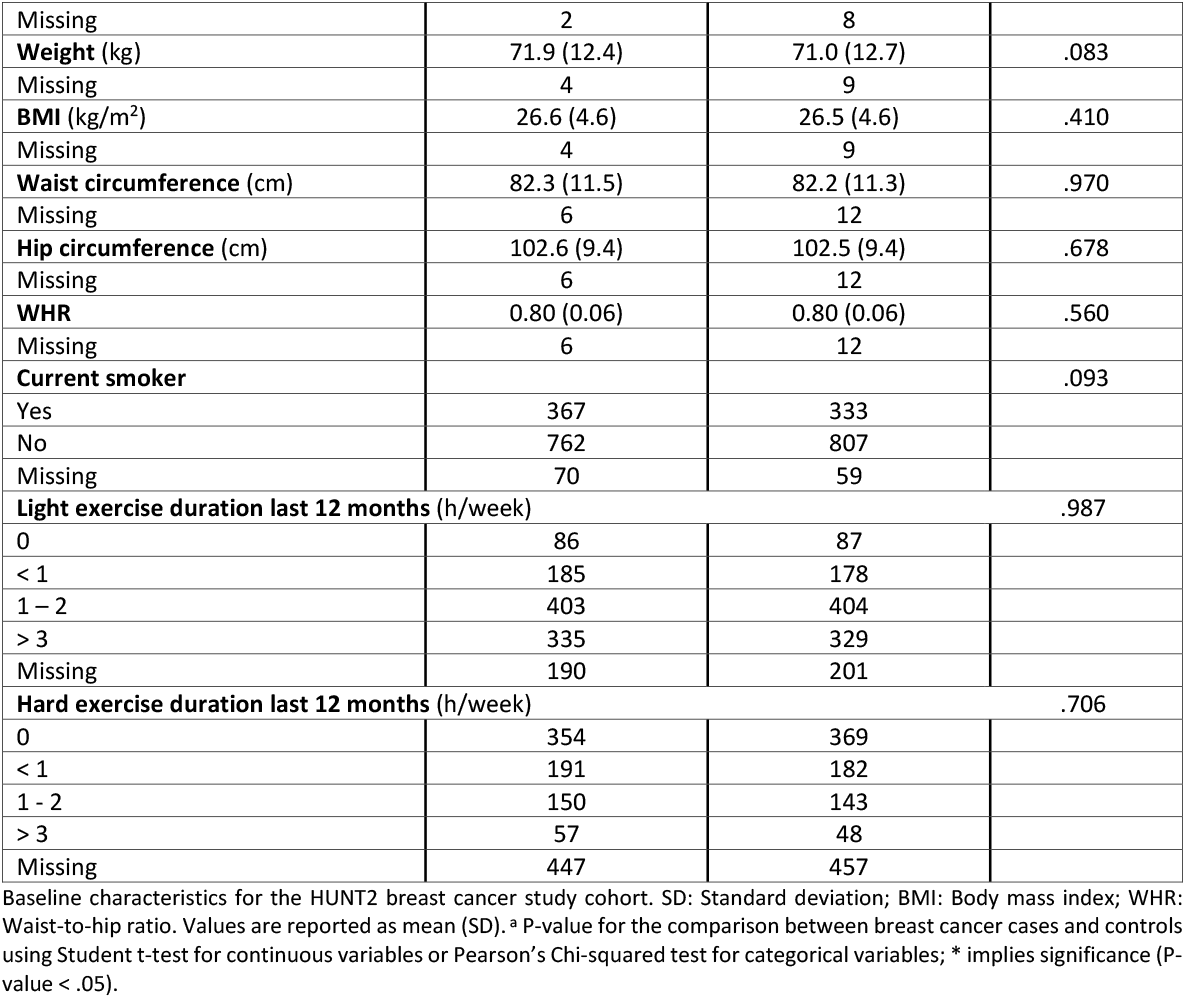
Baseline characteristics for the study cohort.

For breast cancer specific variables (Table 2) approximately 84% of the cancer cases were ER+, 69% were PgR+ and 20% were HER2-, however, this information was missing for 36-44% of the participants, depending on the variable. The majority of the cancers (52%) were stage I tumors, whilst below 4% were advanced (with a distant metastasis). The mean lapse of time between sample collection and the breast cancer diagnosis was 11.7 years (range 0-22 years) and the mean age at diagnosis was 64.4 years.

**Table 2.** Characteristics of breast cancer specific variables for the cases.

| Variable | Mean (SD) or Counts |
| --- | --- |
| **Length of follow-up from blood collection (years)** | 11.7 (6.1) |
| **Age at diagnosis (years)** | 64.4 (13.3) |
| **ER status\*** |  |
| Positive | 639 |
| Negative | 123 |
| Missing | 437 |
| **PgR status** |  |
| Positive | 527 |
| Negative | 233 |
| Missing | 439 |
| **HER2 status** |  |
| Positive | 132 |
| Negative | 538 |
| Missing | 529 |
| **Local metastasis** | 362 |
| **Distant metastasis** | 33 |
| **Tumor size (mm)** | 17 (0.9-120.0)** |
| **Stage (TNM classification)** |  |
| I | 499 |
| II | 331 |
| III | 90 |
| IV | 35 |
| Unknown | 119 |
| **Detection method** |  |
| At screening | 372 |
| Interval cancer | 87 |
| Outside the screening program | 740 |

Baseline characteristics of breast cancer specific variables for the cases in the HUNT2 breast cancer study cohort. SD: Standard deviation; ER: Estrogen receptor; PgR: Progesterone receptor; HER2: Human epidermal growth factor receptor 2; TNM: Tumor, nodes and metastases. \*The cut-off value for defining a breast tumor ER- was changed in 2010 from <10% to <1% in Norway. \*\*Median and range

High correlations were observed between the variables, especially among the lipoprotein subfractions (Figure S2). In general, lipoprotein subfractions from the same lipoprotein main fractions were highly correlated with each other, while weak correlations were observed between some of the lipoprotein subfractions and metabolites.

### Lipoprotein subfractions associated with breast cancer risk in premenopausal women

From the full study cohort, 554 cases were classified as premenopausal and 645 as postmenopausal at inclusion into HUNT2. Postmenopausal women had significantly different lipid profiles compared to premenopausal women, with elevated levels of most lipoprotein subfraction, except for HDL-3 and HDL-4 cholesterol and phospholipids in postmenopausal women (Figure 1, Table S3).

**Figure 1.**
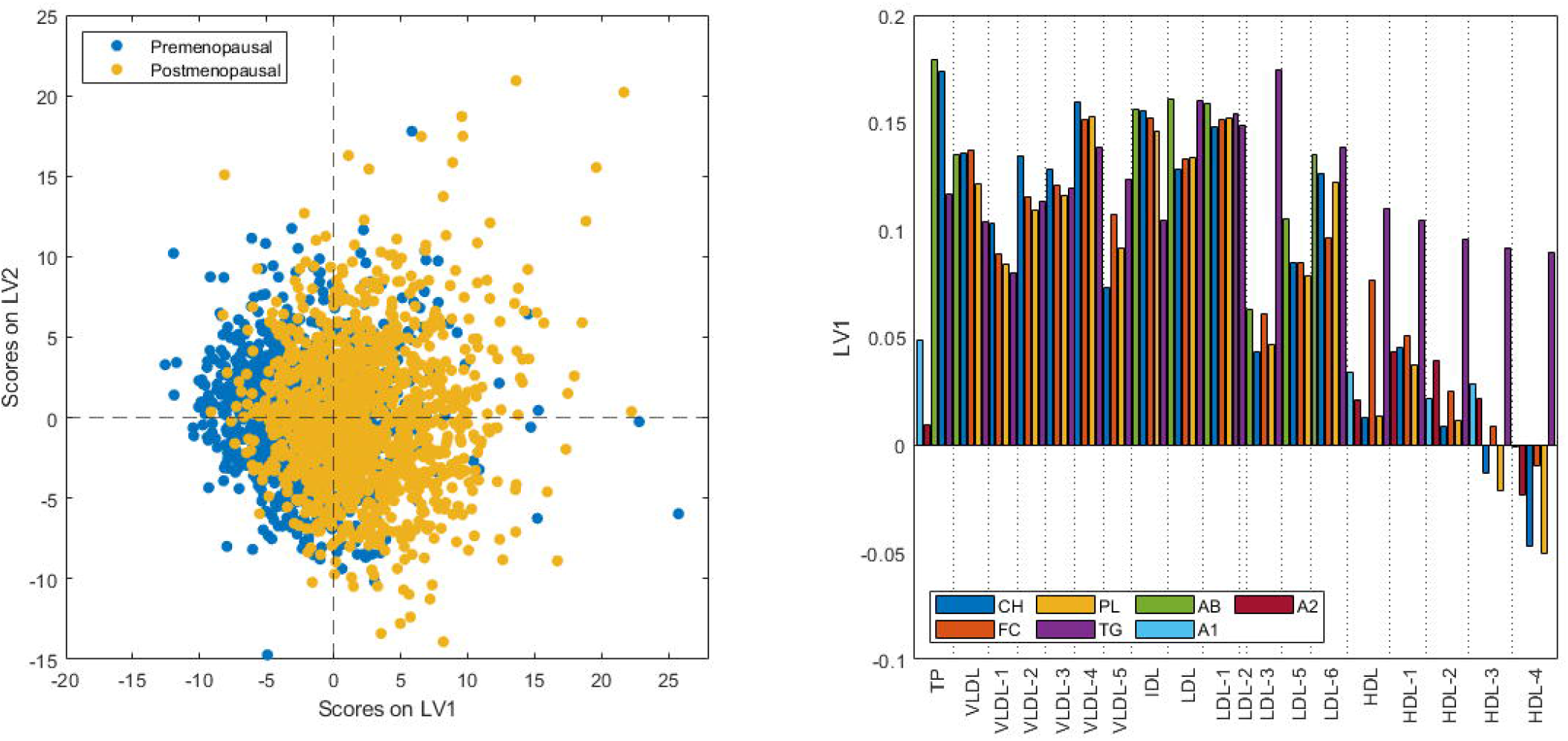
Scores and loading plots from PLS-DA for discrimination of pre- or postmenopausal women from their lipid profiles. The model has been orthogonalized. Prediction accuracy = 70.0% (P-value < .001) with 4 latent variables included. TP: total plasma; VLDL: very-low density lipoprotein; IDL: intermediate-density lipoprotein; LDL: low-density lipoprotein; HDL: high-density lipoprotein; CH: cholesterol; FC: free cholesterol; PL: phospholipids; TG: triglycerides. AB: apolipoprotein-B; A1: apolipoprotein-1; A2: apolipoprotein-2.

For premenopausal women, 38 out of the 89 lipoprotein subfractions had a significant inverse association with long-term breast cancer risk using the baseline model, of which 17 remained significant after correcting for multiple testing (Table 3). In the adjusted model, 14 of the lipoprotein subfractions showed a significant inverse association with long-term breast cancer risk (Figure 2, Table 3). All of the lipoprotein subfractions with a significant association in the adjusted model are VLDL subfractions, except for HDL-4 triglycerides. All associations were in the inverse direction, with odds ratios from 0.77-0.83. Excluding all cases who had reported current use of HRT, or for whom the information about HRT usage was missing, and their matched controls, resulted in similar associations (significant ORs from 0.71-0.80; Table S4). Stratified analysis including ER+ cases or ER-cases only (384 and 72 cases, respectively) showed that the associations were not dependent on the ER status (Table S4). For the postmenopausal women, no significant associations were found between lipoprotein subfractions and breast cancer risk, neither in the full cohort nor in stratified analysis (Results not shown).

**Table 3.** Odds ratios and 95% confidence intervals (CI) per 1 SD increase for lipoprotein subfractions significantly associated with risk of overall breast cancer (p-value < 0.05 and q-value < 0.05) in premenopausal women of the HUNT2 study.

|  |  | Baseline model* |  |  | Adjusted model** |  |  |
| --- | --- | --- | --- | --- | --- | --- | --- |
|  |  | OR (95% CI) | P | P<sub>adj</sub> | OR (95% CI) | P | P<sub>adj</sub> |
| Total plasma | TPA2 | 1.16 (1.01,1.33) | .042 | .107 | 1.12 (0.97,1.29) | .113 | .209 |
|  | **TPTG** | **0.81 (0.69,0.95)** | **.009** | **.046** | 0.81 (0.69,0.96) | .013 | .066 |
| VLDL | **VLAB** | **0.78 (0.67,0.91)** | **.002** | **.044** | **0.77 (0.65,0.90)** | **.002** | **.049** |
|  | **VLCH** | **0.80 (0.69,0.93)** | **.005** | **.046** | **0.80 (0.67,0.94)** | **.007** | **.049** |
|  | **VLFC** | **0.79 (0.67,0.92)** | **.002** | **.044** | **0.78 (0.66,0.92)** | **.003** | **.049** |
|  | **VLPL** | **0.79 (0.68,0.92)** | **.002** | **.044** | **0.78 (0.66,0.91)** | **.002** | **.049** |
|  | VLTG | 0.82 (0.70,0.95) | .009 | .046 | 0.81 (0.69,0.96) | .014 | .066 |
| VLDL-1 | V1CH | 0.84 (0.72,0.98) | .023 | .077 | 0.84 (0.72,0.99) | .042 | .116 |
|  | V1FC | 0.83 (0.72,0.96) | .016 | .061 | 0.83 (0.71,0.98) | .026 | .083 |
|  | V1PL | 0.85 (0.73,0.98) | .026 | .083 | 0.85 (0.72,0.99) | .043 | .117 |
|  | V1TG | 0.86 (0.74,1.00) | .045 | .107 | 0.87 (0.74,1.01) | .076 | .164 |
| VLDL-2 | **V2CH** | **0.79 (0.68,0.92)** | **.003** | **.044** | **0.79 (0.67,0.93)** | **.005** | **.049** |
|  | V2FC | 0.81 (0.69,0.94) | .007 | .046 | 0.81 (0.69,0.95) | .012 | .063 |
|  | **V2PL** | **0.80 (0.69,0.93)** | **.003** | **.044** | **0.80 (0.68,0.93)** | **.005** | **.049** |
|  | **V2TG** | **0.80 (0.69,0.93)** | **.004** | **.044** | **0.80 (0.69,0.94)** | **.006** | **.049** |
| VLDL-3 | V3CH | 0.81 (0.69,0.94) | .007 | .046 | 0.80 (0.68,0.95) | .010 | .057 |
|  | **V3FC** | **0.80 (0.69,0.94)** | **.005** | **.046** | **0.80 (0.67,0.94)** | **.007** | **.049** |
|  | **V3PL** | **0.79 (0.68,0.92)** | **.003** | **.044** | **0.78 (0.67,0.92)** | **.003** | **.049** |
|  | **V3TG** | **0.80 (0.69,0.93)** | **.004** | **.044** | **0.79 (0.68,0.93)** | **.004** | **.049** |
| VLDL-4 | V4CH | 0.82 (0.70,0.96) | .014 | .061 | 0.82 (0.70,0.96) | .015 | .068 |
|  | V4FC | 0.83 (0.71,0.97) | .021 | .074 | 0.82 (0.69,0.97) | .020 | .072 |
|  | **V4PL** | **0.81 (0.69,0.94)** | **.006** | **.046** | **0.80 (0.68,0.93)** | **.005** | **.049** |
|  | **V4TG** | **0.81 (0.69,0.94)** | **.006** | **.046** | **0.80 (0.68,0.93)** | **.004** | **.049** |
| VLDL-5 | V5CH | 0.86 (0.75,0.99) | .040 | .104 | 0.84 (0.73,0.97) | .019 | .072 |
|  | **V5PL** | **0.84 (0.73,0.96)** | **.012** | **.056** | **0.82 (0.71,0.94)** | **.006** | **.049** |
| IDL | IDPL | 0.82 (0.70,0.97) | .018 | .065 | 0.81 (0.68,0.97) | .019 | .072 |
|  | IDTG | 0.83 (0.71,0.96) | .015 | .061 | 0.83 (0.71,0.98) | .026 | .083 |
| LDL | LDTG | 0.83 (0.70,0.96) | .016 | .061 | 0.82 (0.69,0.97) | .018 | .072 |
| LDL-5 | L5AB | 0.86 (0.74,0.99) | .040 | .104 | 0.83 (0.72,0.97) | .017 | .072 |
|  | L5CH | 0.88 (0.76,1.01) | .073 | .133 | 0.85 (0.73,0.98) | .029 | .086 |
|  | L5PL | 0.87 (0.75,1.01) | .061 | .122 | 0.84 (0.72,0.97) | .021 | .073 |
| LDL-6 | L6AB | 0.84 (0.72,0.98) | .028 | .084 | 0.85 (0.72,1.00) | .045 | .118 |
|  | L6TG | 0.84 (0.72,0.98) | .032 | .096 | 0.86 (0.73,1.00) | .056 | .141 |
| HDL | HDTG | 0.85 (0.75,0.96) | .009 | .046 | 0.85 (0.74,0.96) | .011 | .059 |
| HDL-1 | H1TG | 0.86 (0.76,0.98) | .022 | .074 | 0.87 (0.76,0.98) | .028 | .086 |
| HDL-2 | H2TG | 0.88 (0.78,1.00) | .043 | .107 | 0.89 (0.78,1.01) | .064 | .151 |
| HDL-3 | H3CH | 1.15 (1.01,1.30) | .035 | .098 | 1.11 (0.98,1.27) | .113 | .209 |
|  | H3FC | 1.16 (1.01,1.32) | .035 | .098 | 1.13 (0.98,1.29) | .096 | .186 |
| HDL-4 | **H4TG** | **0.85 (0.75,0.96)** | **.012** | **.056** | **0.83 (0.73,0.95)** | **.008** | **.049** |

TP: total plasma; VLDL: very-low density lipoprotein; IDL: intermediate-density lipoprotein; LDL: low-density lipoprotein; HDL: high-density lipoprotein; CH: cholesterol; FC: free cholesterol; PL: phospholipids; TG: triglycerides. AB: apolipoprotein-B; A2: apolipoprotein-2.
\*Baseline model: adjusted for lab for NMR analyses and matching variable (age at participation in the HUNT2 study)
\*\*Adjusted model: in addition to lab and age, this model is adjusted for no. of full-term pregnancies, age at menarche, BMI, alcohol consumption and smoking status. Bold font indicates variables significant in the adjusted model after multiple testing correction.

**Figure 2.**
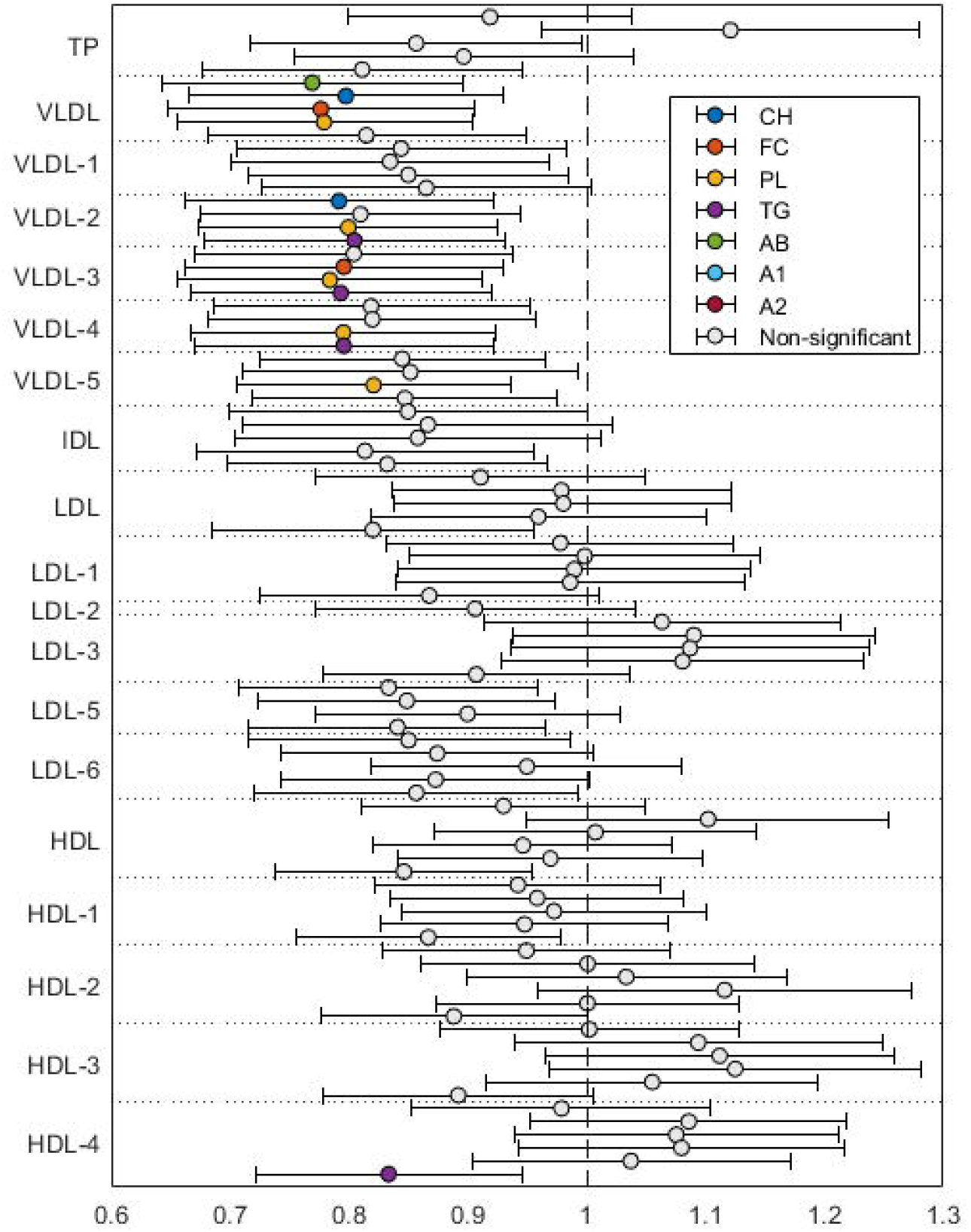
Odds ratio per SD for lipoprotein subfractions associated with long-term breast cancer risk in premenopausal women participating in the HUNT2 study. Colored subfractions indicate subfractions significantly associated with breast cancer risk in the fully adjusted model, after correction for multiple testing. Each color represents a property of the subfraction. TP: total plasma; VLDL: very-low density lipoprotein; IDL: intermediate-density lipoprotein; LDL: low-density lipoprotein; HDL: high-density lipoprotein; CH: cholesterol; FC: free cholesterol; PL: phospholipids; TG: triglycerides. AB: apolipoprotein-B; A1: apolipoprotein-1; A2: apolipoprotein-2.

### Circulating metabolites

Analyses performed on premenopausal women showed a significant positive association between acetate and breast cancer risk in the baseline model (P-value = .037). In the adjusted model, acetate and phenylalanine showed a significant positive association with breast cancer risk (P-values <.05; Table S5). However, none of the associations remained significant after correcting for multiple testing. Similarly as for lipoprotein subfractions, no significant associations were found between circulating metabolites and breast cancer risk for postmenopausal women.

### Prediction of a future cancer from prediagnostic serum metabolic profiles

A weak but significant discrimination between breast cancer cases and controls was obtained for premenopausal women (Accuracy = 53%; P-value = .028; Table 4) from lipoprotein subfractions. Adding information on established breast cancer risk factors did not increase the prediction accuracy (Accuracy = 53%; P-value = .032; Table 4). No significant discrimination was obtained for postmenopausal women.

**Table 4.** PLS-DA prediction models for predicting a future breast cancer from prediagnostic levels of lipoprotein subfractions.

|  | Follow-up period | n | Accuracy | LV | P<sub>perm</sub> |
| --- | --- | --- | --- | --- | --- |
| Premenopausal women | Complete | 1108 | 53 % | 1 | .027 |
|  | Complete * | 1108 | 53 % | 1 | .021 |
|  | 15 years | 596 | 51 % | 1 | .403 |
|  | 10 years | 310 | 51 % | 6 | .405 |
|  | 5 years | 120 | 52 % | 1 | .392 |
| Postmenopausal women | Complete | 1290 | 51 % | 1 | .300 |
|  | 15 years | 950 | 50 % | 1 | .482 |
|  | 10 years | 652 | 51 % | 1 | .280 |
|  | 5 years | 288 | 50 % | 3 | .644 |

\* : including clinical variables (lab, age, no. of full-term pregnancies, age at menarche, BMI, alcohol consumption, smoking status and current use of HRT) in addition to lipoprotein subfractions. n: total number of subjects; P<sub>perm</sub>: P-value from permutation testing (1000 permutations); LV: latent variable.

### Discussion

The discovery of novel biomarkers for early breast cancer development has several clinical applications, such as insight into metabolic pathways that may represent new therapeutic targets, and early identification of individuals eligible for primary prevention. In this study we analyzed the association of circulating lipoprotein subfractions and metabolites with breast cancer incidence, and assessed the predictive value of serum metabolic profiles of healthy females. We found significant associations between multiple circulating lipoprotein subfractions and breast cancer risk 0-22 years after blood collection. This study is the first to report associations between lipoprotein subfractions and long-term breast cancer risk.

Our results reveal alterations in the lipid metabolism of premenopausal women many years before they develop breast cancer. We found that high levels of circulating cholesterol, free cholesterol, phospholipids, and triglycerides in VLDL subfractions have a protective effect from developing breast cancer, even when adjusting for clinical risk factors including lifestyle factors, however only for premenopausal women. More specifically, several VLDL2-4 subfractions were inversely associated with breast cancer risk. VLDLs are large particles consisting mainly of triglycerides. Their size depends on the content of triglycerides while the rate of synthesis depends on the availability of triglycerides (Figure 3) [25].

**Figure 3.**
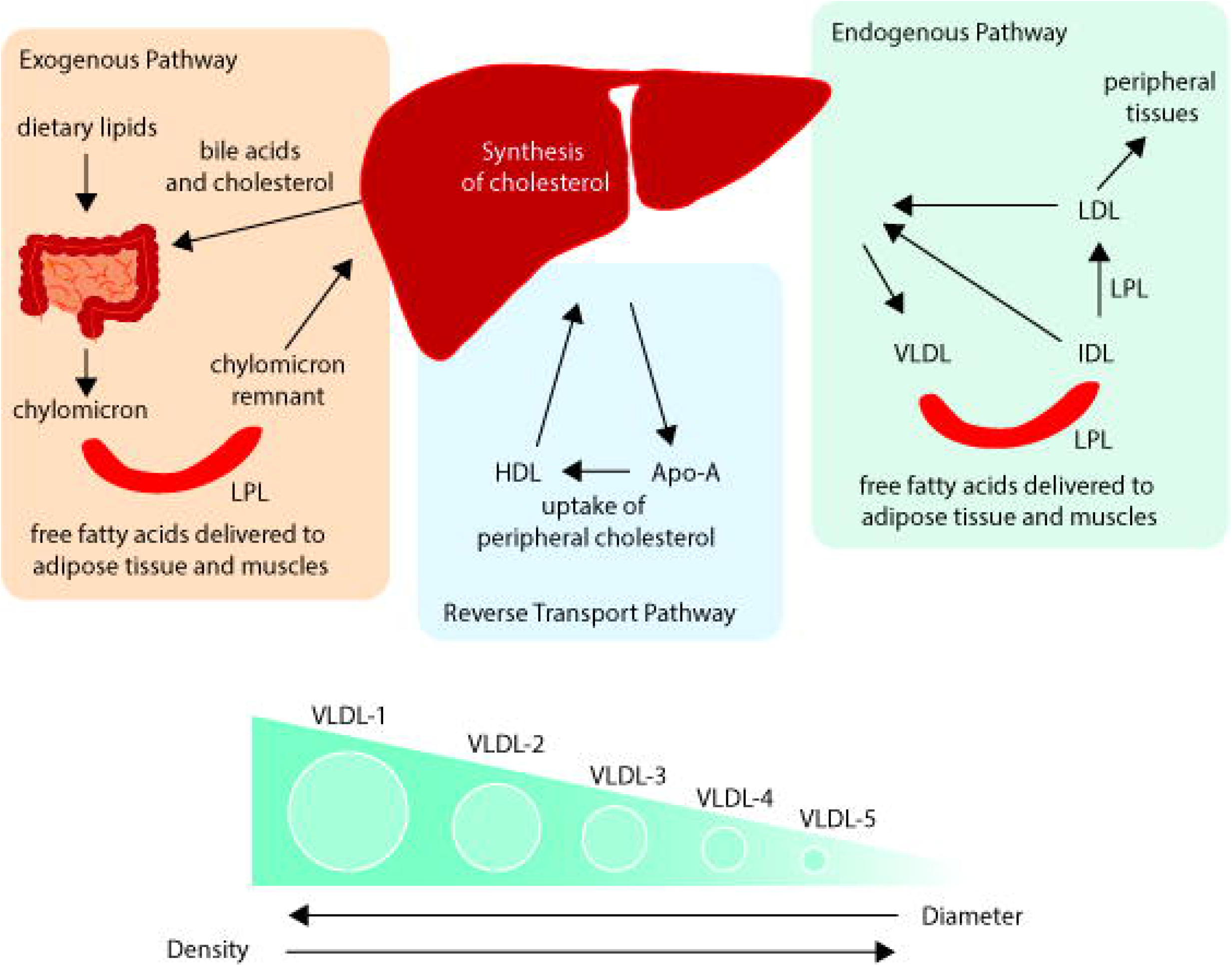
Lipoprotein metabolism. Exogeneous pathway: The exogeneous pathway starts in the intestine, where dietary lipids are hydrolyzed. The lipids are then assembled to chylomicrons, which are transported through the bloodstream to the liver. In this process free fatty acids are delivered to adipose tissue and muscles through the enzyme lipoprotein lipase (LPL), for energy and storage. Endogeneous pathway: VLDL is synthesized by the liver and transported through the bloodstream. There free fatty acids are delivered to adipose tissue and muscles through LPL, and VLDLs are reduced to IDLs and finally to LDLs as fatty acids are cleaved off. These are taken up by the liver, however a residual fraction of LDLs turn into foam cells and may form atherosclerotic plaque [21]. The reverse transport pathway: HDL is synthesized in the liver and enters the bloodstream, where it removes cholesterol from peripheral tissue, through the action of Apo-A which acts as an acceptor, and transfers it back to the liver. Lipoprotein subfractions are numbered according to increasing density, as illustrated for VLDL in the figure.

Other studies, utilizing traditional clinical chemistry methods to assess serum lipids have suggested that levels of triglycerides may be inversely associated with breast cancer risk. No study to date has looked at triglycerides or VLDL levels and premenopausal breast cancer risk, however a recent study by Bendinelli et al. investigated associations between VLDL subfractions and high mammographic breast density [26], which is a strong risk factor for breast cancer. They found free cholesterol, triglycerides, cholesterol and Apo-B levels in VLDL main fraction, and the subfractions VLDL-1 and VLDL-5 to be inversely associated with high mammographic breast density, supporting our finding of inverse associations between VLDL parameters and breast cancer risk. Similarly, a study on NMR metabolic profiles found an inverse association between lipids and lipoproteins and increased breast cancer risk, however, this study did not include subfraction analysis or stratified analysis based on the menopausal status [13].

Studies have shown that circulating estrogens are positively associated with breast cancer risk [27], especially in postmenopausal women [28-30]. Estrogen levels play an important role in the regulation of lipid metabolism, and are shown to be negatively correlated with triglycerides and VLDLs [31, 32]. Impaired estrogen signaling is associated with the development of metabolic disorders, and an estrogen deficiency will lead to insulin resistance, which in turn will cause increased lipogenesis, triglyceride accumulation and increased VLDL production in the liver [33]. In our study we found that in addition to VLDL subfractions, total triglycerides, IDL, LDL and HDL 2-4 triglycerides are inversely associated with premenopausal breast cancer risk. The protective effect of VLDL subfractions and triglycerides for premenopausal breast cancer risk observed in this study might thus reflect hormonal activity. Sensitivity analysis showed that these associations were not dependent on the ER status of the tumor or the use of HRT.

Several LDL 5-6 subfractions were inversely associated with breast cancer (Apo-B in LDL 5-6, phospholipids and cholesterol in LDL-5 and triglycerides in LDL-6), although these associations did not reach statistical significance when correcting for multiple testing. Interestingly, no association was found between LDL-1 or LDL-3 and breast cancer risk. As illustrated in Figure 3, LDL is synthesized in the same lipid pathway as VLDL, and in general smaller LDLs are the most atherogenic [34], thus these findings are surprising. However, due to the presence of a contamination in the serum samples (Figure S1, Supplementary methods), most parameters from LDL-2 (except triglycerides) and all parameters from LDL-4 subfractions were removed, and the associations of these parameters and breast cancer risk could not be assessed. Collectively, our results suggest that early breast cancer formation in premenopausal women is likely driven by hormonal activity rather than an unhealthy lifestyle. This should thus be further elucidated.

We found no significant associations between lipoprotein subfractions and long-term breast cancer risk for postmenopausal women. This is in accordance with a recent study by Jobard et al. showing that a perturbed metabolism is associated with increased breast cancer risk in premenopausal women only [35]. Furthermore, our finding of different lipid profiles between pre- and postmenopausal women is in accordance with previous studies which have shown that the lipid profile is highly dependent on the menopausal status [36-38], reflected in significantly higher cholesterol and total, LDL and VLDL (but not HDL) triglyceride levels in postmenopausal women. Other studies have shown that a weight gain or high BMI is associated with increased breast cancer risk in postmenopausal women, while the opposite is true for premenopausal women [39, 40], thus the lipid metabolism is clearly affected by hormone activities.

Although several metabolites significantly associated with breast cancer risk have been reported in other studies [12-16, 35, 41-43], there is a heterogeneity in the analytical platforms used and type of biological medium. Three recent studies have reported serum metabolic alterations, measured by NMR, associated with overall breast cancer risk [13] or premenopausal breast cancer risk [17, 43], with metabolic panels that overlap with ours. We found weak associations only between acetate and phenylalanine, and premenopausal breast cancer risk, which have not been reported in previous studies.

This study presents one of the largest prospective analysis of serum metabolic profiles and breast cancer risk to date. The large study cohort, long follow-up period and availability of numerous lifestyle factors allowed for evaluating the behavior of significant associations when adjusting for established breast cancer risk factors. Associations between metabolic factors and breast cancer risk are in general modest, in terms of their odds ratios, as compared to other diseases such as diabetes, where metabolomics research findings have been replicated several times [44, 45]. This implies a lack of accurate predictive value of serum metabolic profiles of healthy females and breast cancer, which we have observed in this study.

In conclusion, we identified several associations between lipoprotein subfractions and long-term risk of breast cancer in premenopausal women. In particular, we found inverse associations in several VLDL subfractions and breast cancer, revealing an altered metabolism in the endogenous lipid pathway many years prior to a breast cancer diagnosis.

## Data Availability

To protect participants privacy, HUNT Research Centre aims to limit storage of data outside HUNT databank, and cannot deposit data in open repositories. HUNT databank has precise information on all data exported to different projects and are able to reproduce these on request. There are no restrictions regarding data export given approval of applications to HUNT Research Centre. For more information see: http://www.ntnu.edu/hunt/data.

## Notes

### Author contributions

Conceptualization: JD, TFB, GFG. Data curation: JD, TFB, GFG. Formal analysis: JD, TA, TFB, GFG. Funding acquisition: TFB, GFG. Investigation: JD, HS, TA, FW, FF, CC, GFG. Methodology: JD, HS, TA, MS, TFB, GFG. Software: HS, MS. Supervision: TFB, GFG. Visualization: JD. Writing – original draft: JD. Writing – review & editing: JD, HS, TA, FW, FF, CC, MS, TFB, GFG.

## Acknowledgements

The Trøndelag Health Study (HUNT) is a collaboration between HUNT Research Centre (Faculty of Medicine and Health Sciences, Norwegian University of Science and Technology NTNU), Trøndelag County Council, Central Norway Regional Health Authority, and the Norwegian Institute of Public Health. The NMR analyses were performed at the MR Core Facility, Norwegian University of Science and Technology (NTNU), funded by the Faculty of Medicine at NTNU and Central Norway Regional Health Authority, and at Bruker BioSpin GmbH, Germany.

## Data availability statement

The Trøndelag Health Study (HUNT) has invited persons aged 13 - 100 years to four surveys between 1984 and 2019. Comprehensive data from more than 140,000 persons having participated at least once and biological material from 78,000 persons are collected. The data are stored in HUNT databank and biological material in HUNT biobank. HUNT Research Centre has permission from the Norwegian Data Inspectorate to store and handle these data. The key identification in the data base is the personal identification number given to all Norwegians at birth or immigration, whilst de-identified data are sent to researchers upon approval of a research protocol by the Regional Ethical Committee and HUNT Research Centre. To protect participants’ privacy, HUNT Research Centre aims to limit storage of data outside HUNT databank, and cannot deposit data in open repositories. HUNT databank has precise information on all data exported to different projects and are able to reproduce these on request. There are no restrictions regarding data export given approval of applications to HUNT Research Centre. For more information see: http://www.ntnu.edu/hunt/data.

