## Supplemental Materials for "Lipoprotein and metabolite associations to breast cancer risk in the HUNT2 study"

### Supplementary Material

#### Supplementary Methods

##### NMR Experiments and Data Preprocessing

Serum samples were collected in the years 1995-97 and were stored in -80°C until analysis. After thawing at room temperature, 150 µL serum was mixed with 150 µL buffer [$D_2O$ (20% in $H_2O$) with 0.075 M $Na_2HPO_4$, 6 mM $NaN_3$, 4.6 mM 3-(trimethylsilyl)-2,2,3,3-tetradeuteropropanoic acid (TSP-$d_4$), pH 7.4], and transferred to 3 mm NMR tubes. Quality control (QC) samples were prepared from pooled serum samples of 10 anonymous donors from the Norwegian blood bank (46 samples in total). One QC sample per 60 HUNT sample was run for the NMR analysis, to assess the quality of the NMR acquisitions and identify instrumental drifts, and all samples were analyzed in a random order. Approximately half of the serum samples were analyzed locally at the MR Core facility, NTNU, in Trondheim, while the second half was shipped to Bruker BioSpin GmbH, Germany, for analyses. NMR analyses were carried out on a Bruker Avance III HD Ultrashield Plus 600 MHz spectrometer (Bruker BioSpin) equipped with a 5 mm TCI probe in Trondheim, and an Avance-IVDr spectrometer (Bruker BioSpin) at the lab in Germany. The equipment at both labs has been calibrated for use of the same protocol. Sample handling and data acquisition were automatically performed using SampleJet sample changer and Icon-NMR on Topspin 3.5 (Bruker BioSpin). NMR spectra were recorded using one-dimensional nuclear Overhauser effect spectroscopy (1D-NOESY) and Carr-Pucell-Meiboom-Gill (CPMG) experiments. Both experiments were performed at 310 K and applied irradiation (25 Hz) on the water resonance during relaxation delay (4 s) and mixing time (10 ms). The 1D-NOESY experiment applied pulse sequence "noesygpprd" (Bruker nomenclature) using 96k data points and 30 ppm spectral width. 32 scans were recorded, and the free induction decays were Fourier-transformed after zero filling (128k real data points) and 0.3 Hz line broadening. The CPMG experiment (pulse sequence "cpmgpr1d", Bruker nomenclature) was recorded with 72k data points, 20 ppm spectral width and 32 scans. Data was zero filled to 128k data points, line broadening (0.3 Hz) before Fourier-transformation. The study cohort included 1199 cases and 1199 matched controls.

##### Metabolite quantification

Small-molecular metabolites were quantified from CPMG spectra to be able to quantify more metabolites than provided by B.I.QUANT-PS from NOESY spectra. $T_2$ attenuation in CPMG was compensated for by additional $T_2$ measurements. CPMG spectral data were transferred to Matlab R2020a for preprocessing. Spectra of samples run at NTNU and at Bruker BioSpin were preprocessed separately due to differences in peak positions of individual metabolites. The left peak of the alanine doublet at 1.47 ppm was used as a chemical shift reference for an initial alignment of the spectra, followed by a more thorough peak alignment using the icoshift function, where the mean spectra was used as the reference [1]. The spectral region 0.2 to 9.2 ppm was chosen as the region of interest. Spectral peaks were assigned to metabolites using the human metabolome database (HMDB), published literature, an in-house overview over previously assigned spectral peaks in serum based on 2D HSQC acquisitions and the STOSCY algorithm [2]. Areas under the spectral peaks were integrated, and corrected for the number of protons giving rise to the signals. Thereafter, peaks were adjusted for $T_2$ relaxation times (Table S1). Transverse magnetization decay is given by $M_x(t) = M_x(0)e^{-R_{xy}t}$, where $M_x$ is the magnetization at time $t$ and $R_{xy}$ is the transverse relaxation rate constant [3]. $T_2$ is the reciprocal of the rate constant: $T_2 = 1/R_{xy}$, and can be measured with a spin-echo sequence. By running an experiment using a short pulse to form multiple echoes, the decay can be observed and used to estimate $T_2$ values [4]. To obtain $T_2$ relaxation times, we performed CMPG experiments on three separate serum samples, and modelled the exponential decay for each signal separately, based on the area under the signal. The exponential decay was modelled using a two-component exponential function, yielding a separate component for the decay of the lipid signals and the metabolite decay. To obtain a better fit of the exponential function to the data, which tended to overestimate the decay, every second point at the beginning of the decay was left out when fitting the line, giving more weight to the smaller value closer to zero. The $T_2$ values varied across these three samples, due to their slightly different metabolic compositions. Therefore, the signal (spectral integral) was corrected based on the mean value of the $T_2$ values of the three serum samples. Peaks arising from the same metabolite were averaged, giving a total of 28 quantified metabolites. The concentration of glucose was set equal to the automatically quantified glucose concentration (Bruker B.I. Quant-PS™) [5] and the remaining metabolite concentrations were scaled accordingly using the same factor, thus giving absolute metabolite concentrations.

#### Lipoprotein Parameter Analysis

Lipoprotein parameters were automatically quantified using Bruker IVDr Lipoprotein Subclass Analysis (B.I.LISA™) software, from Bruker BioSpin. This method utilizes the broad lipid signals from the methyl ($-CH_3$) groups at 0.85 ppm and methylene ($-CH_2-$) groups at 1.57 ppm of the NMR spectrum, to provide a detailed picture of circulating lipoproteins [6]. Concentrations of lipids [cholesterol (CH), free cholesterol (FC), triglycerides (TG), and phospholipids (PL)] in total serum and in four main lipoprotein classes: very low-, intermediate-, low-, and high-density lipoproteins (VLDL, IDL, LDL, and HDL) and 15 subclasses (VLDL

1−5, LDL 1−6, and HDL 1−4) were provided. In addition, serum levels of apolipoproteins (Apo-A1, Apo-A2, and Apo-B) in the lipoproteins, 12 calculated parameters (ratios of LDL-CH/HDL-CH and Apo- B/Apo-A1, and 10 particle numbers (total serum, VLDL, IDL, LDL, and LDL 1−6) were provided, giving a total of 112 lipoprotein parameters. Particle number variables, highly correlated with Apo-B quantifications (ϱ=1), and calculated rations were considered redundant and removed from the variable list.

NMR analysis revealed a signal interfering with the broad lipid peak at 0.85 ppm. The contamination was identified as neopentyl glycol from HSQC and HMBC experiments. A spiking experiment was thus conducted, where serum samples from 9 healthy donors were spiked with known concentrations of neopentyl glycol. Spectra corresponding to the spiked spectra were compared to spectra of clean serum from the same donors. These spectra were sent to Bruker BioSpin, which developed a method for removing the peak arising from the contamination before lipoprotein quantification by B.I.LISA™. This method was then applied to all spectra of the HUNT samples, and Supplemental Figure S1 shows the results of this approach for two randomly selected donors. To assess the success of this adjustment, percentage changes between the lipoprotein parameters of the clean and the spiked samples with the peak removed, were calculated. Based on the spiking experiment, which included 9 donors, 3 of the variables had a percentage difference exceeding +/- 10% for 9 of the donors. Four of the variables had a percentage difference exceeding +/- 10% for 8 of the donors, 4 variables for 7 of the donors, and 7 variables for 6 and 5 of the donors. One of the donors was excluded due to a bad fit and hence consistently large percentage differences. Parameters with a percentage change exceeding 20% for 2 of the 9 donors were excluded from the analysis, resulting in 99 lipoprotein variables for data analysis. Variables excluded were L2PN, L4PN, L4TG, L5TG, L2CH, L4CH, L2FC, L4FC, L2PL, L4PL, L2AB, L4AB and H1A1.

#### Supplementary Tables

**Table S1.** Overview over estimated T2 values for signals corresponding to metabolites included in this study, and the corresponding coefficients of variation (CVs) calculated from quality control samples.

| Metabolite | Chemical shift (peak multiplicity) | T2-values (ms) | CV (%) |
|---|---|---|---|
| Phenylalanine | 7. 34 (d); 7.43 (t); 7.38 (t) | 0.41; 0.60; 0.58 | 11.6 |
| Histidine | 7.72 (s); 7.06 (s) | 1.42; 1.44 | 16.1 |
| Formate | 8.46 (s) | 5.55 | 29.8 |
| Glucose | 5.24 (d); 3.9 (q); 3.53 (q); 3.35 (q) | 1.23; 0.58; 1.32; 1.06 | 8.4 |
| Tyrosine | 6.9 (d); 7.2 (d) | 0.74; 0.50 | 13.1 |
| Creatine | 2.93 (s) | 0.38 | 7.8 |
| Creatinine | 4.05 (s); 3.04 (s) | 0.42; 1.12 | 9.3 |
| Lactate | 4.11 (q) | 1.31 | 7.4 |
| Valine | 3.6 (d); 0.98 (d) 1.03 (d) | 1.60; 0.85; 1.01 | 8.5 |
| Glycine | 3.56 (s) | 0.78 | 13.3 |
| Methanol | 3.36 (s) | 2.59 | 18.7 |
| Proline betaine | 3.1 (s); 3.29 (s) | 0.38; 0.45 | 6.3 |
| Dimethyl-sulfone | 3.15 (s) | 2.50 | 16.2 |
| Lysine | 3.02 (d); 1.70 (m) | 0.13; 0.12 | 4.6 |
| Ornithine | 3.06 (d) | 2.97 | 5.5 |
| Methionine | 2.63 (d) | 0.19 | 11.7 |
| Glutamine | 2.45 (m) | 0.53 | 9.5 |
| Citrate | 2.53 (d) | 0.73 | 21.6 |
| Acetate | 1.90 (s) | 1.11 | 8.0 |
| Acetoacetate | 2.27 (s) | 0.18 | 10.4 |
| 3-Hydroxybutyrate | 2.30 (q); 2. 39 (q); 1.19 (d) | 0.10; 0.34; 0.10 | 8.2 |
| Glutamate | 2.35 (m) | 0.22 | 5.3 |
| Pyruvate | 2.36 (s) | 0.66 | 9.4 |
| Alanine | 1.47 (d) | 0.70 | 8.2 |
| Ethanol | 1.16 (t) | 0.09 | 14.6 |
| Isoleucine | 0.10 (d) | 0.34 | 12.0 |
| 2-Methylglutarate | 1.06 (d) | 0.12 | 14.3 |
| Leucine | 0.95 (t) | 0.46 | 11.5 |

s: singlet; d: doublet; t: triplet; q: quartet; m:multiplet.

**Table S2.** Coefficients of variation (CVs) of lipoprotein subfractions included in this study. CVs were calculated from quality control samples.

| Lipoprotein Parameter | | CV (%) |
|---|---|---|
| **Main Parameters** | | |
| TPTG | Triglycerides [mg/dL] | 3.9 |
| TPCH | Cholesterol [mg/dL] | 4.8 |
| LDCH | LDL Cholesterol, [mg/dL] | 5.4 |
| HDCH | HDL Cholesterol, [mg/dL] | 3.1 |
| TPA1 | Apo-A1, [mg/dL] | 4.7 |
| TPA2 | Apo-A2, [mg/dL] | 6.3 |
| TPAB | Apo-B100, [mg/dL] | 3.4 |
| **Calculated Figures** | | |
| LDHD | LDL Cholesterol / HDL | 3.5 |
| ABA1 | Apo-A1 / Apo-B100, [-/-] | 2.4 |
| TBPN | Total ApoB Particle Number, | 3.4 |
| VLPN | VLDL Particle Number, [nmol/L] | 3.2 |
| IDPN | IDL Particle Number, [nmol/L] | 16.8 |
| LDPN | LDL Particle Number, [nmol/L] | 3.8 |
| L1PN | LDL-1 Particle Number, [nmol/L] | 7.6 |
| L2PN * | LDL-2 Particle Number, [nmol/L] | 13.7 |
| L3PN | LDL-3 Particle Number, [nmol/L] | 18.3 |
| L4PN * | LDL-4 Particle Number, [nmol/L] | 14.6 |
| L5PN | LDL-5 Particle Number, [nmol/L] | 10.0 |
| L6PN | LDL-6 Particle Number, [nmol/L] | 15.3 |
| **Lipoprotein Main Fractions** | | |
| VLTG | Triglycerides, VLDL [mg/dL] | 2.9 |
| IDTG | Triglycerides, IDL [mg/dL] | 6.7 |
| LDTG | Triglycerides, LDL [mg/dL] | 10.3 |
| HDTG | Triglycerides, HDL [mg/dL] | 10.1 |
| VLCH | Cholesterol, VLDL [mg/dL] | 7.1 |
| IDCH | Cholesterol, IDL [mg/dL] | 20.5 |
| VLFC | Free Cholesterol, VLDL [mg/dL] | 2.6 |
| IDFC | Free Cholesterol, IDL [mg/dL] | 21.3 |
| LDFC | Free Cholesterol, LDL [mg/dL] | 7.0 |
| HDFC | Free Cholesterol, HDL [mg/dL] | 8.6 |
| VLPL | Phospholipids, VLDL [mg/dL] | 2.8 |
| IDPL | Phospholipids, IDL [mg/dL] | 15.8 |
| LDPL | Phospholipids, LDL [mg/dL] | 5.0 |
| HDPL | Phospholipids, HDL [mg/dL] | 5.4 |
| HDA1 | Apo-A1, HDL [mg/dL] | 4.4 |
| HDA2 | Apo-A2, HDL [mg/dL] | 5.5 |
| VLAB | Apo-B, VLDL [mg/dL] | 3.2 |
| IDAB | Apo-B, IDL [mg/dL] | 16.8 |
| LDAB | Apo-B, LDL [mg/dL] | 3.8 |
| **VLDL Subfractions** | | |
| V1TG | Triglycerides, VLDL-1 [mg/dL] | 3.4 |
| V2TG | Triglycerides, VLDL-2 [mg/dL] | 6.2 |
| V3TG | Triglycerides, VLDL-3 [mg/dL] | 9.9 |
| V4TG | Triglycerides, VLDL-4 [mg/dL] | 6.8 |
| V5TG | Triglycerides, VLDL-5 [mg/dL] | 9.9 |

| | | |
|---|---|---|
| V1CH | Cholesterol, VLDL-1 [mg/dL] | 4.4 |
| V2CH | Cholesterol, VLDL-2 [mg/dL] | 9.3 |
| V3CH | Cholesterol, VLDL-3 [mg/dL] | 21.0 |
| V4CH | Cholesterol, VLDL-4 [mg/dL] | 11.7 |
| V5CH | Cholesterol, VLDL-5 [mg/dL] | 15.6 |
| V1FC | Free Cholesterol, VLDL-1 [mg/dL] | 6.9 |
| V2FC | Free Cholesterol, VLDL-2 [mg/dL] | 10.9 |
| V3FC | Free Cholesterol, VLDL-3 [mg/dL] | 15.1 |
| V4FC | Free Cholesterol, VLDL-4 [mg/dL] | 16.1 |
| V5FC | Free Cholesterol, VLDL-5 [mg/dL] | 29.8 |
| V1PL | Phospholipids, VLDL-1 [mg/dL] | 4.3 |
| V2PL | Phospholipids, VLDL-2 [mg/dL] | 8.0 |
| V3PL | Phospholipids, VLDL-3 [mg/dL] | 16.0 |
| V4PL | Phospholipids, VLDL-4 [mg/dL] | 8.2 |
| V5PL | Phospholipids, VLDL-5 [mg/dL] | 9.3 |
| **LDL Subfractions** | | |
| L1TG | Triglycerides, LDL-1 [mg/dL] | 8.5 |
| L2TG | Triglycerides, LDL-2 [mg/dL] | 18.9 |
| L3TG | Triglycerides, LDL-3 [mg/dL] | 4.4 |
| L4TG * | Triglycerides, LDL-4 [mg/dL] | 19.2 |
| L5TG * | Triglycerides, LDL-5 [mg/dL] | 13.4 |
| L6TG | Triglycerides, LDL-6 [mg/dL] | 11.4 |
| L1CH | Cholesterol, LDL-1 [mg/dL] | 8.1 |
| L2CH * | Cholesterol, LDL-2 [mg/dL] | 14.0 |
| L3CH | Cholesterol, LDL-3 [mg/dL] | 18.5 |
| L4CH * | Cholesterol, LDL-4 [mg/dL] | 11.7 |
| L5CH | Cholesterol, LDL-5 [mg/dL] | 10.0 |
| L6CH | Cholesterol, LDL-6 [mg/dL] | 16.6 |
| L1FC | Free Cholesterol, LDL-1 [mg/dL] | 11.7 |
| L2FC * | Free Cholesterol, LDL-2 [mg/dL] | 13.1 |
| L3FC | Free Cholesterol, LDL-3 [mg/dL] | 14.3 |
| L4FC * | Free Cholesterol, LDL-4 [mg/dL] | 11.0 |
| L5FC | Free Cholesterol, LDL-5 [mg/dL] | 7.5 |
| L6FC | Free Cholesterol, LDL-6 [mg/dL] | 6.5 |
| L1PL | Phospholipids, LDL-1 [mg/dL] | 7.5 |
| L2PL * | Phospholipids, LDL-2 [mg/dL] | 13.2 |
| L3PL | Phospholipids, LDL-3 [mg/dL] | 17.1 |
| L4PL * | Phospholipids, LDL-4 [mg/dL] | 11.3 |
| L5PL | Phospholipids, LDL-5 [mg/dL] | 8.6 |
| L6PL | Phospholipids, LDL-6 [mg/dL] | 13.5 |
| L1AB | Apo-B, LDL-1 [mg/dL] | 7.6 |
| L2AB * | Apo-B, LDL-2 [mg/dL] | 13.7 |
| L3AB | Apo-B, LDL-3 [mg/dL] | 18.3 |
| L4AB * | Apo-B, LDL-4 [mg/dL] | 14.6 |
| L5AB | Apo-B, LDL-5 [mg/dL] | 10.0 |
| L6AB | Apo-B, LDL-6 [mg/dL] | 15.3 |
| **HDL Subfractions** | | |
| H1TG | Triglycerides, HDL-1 [mg/dL] | 19.9 |
| H2TG | Triglycerides, HDL-2 [mg/dL] | 17.3 |
| H3TG | Triglycerides, HDL-3 [mg/dL] | 10.6 |
| H4TG | Triglycerides, HDL-4 [mg/dL] | 2.8 |

| | | |
|---|---|---|
| H1CH | Cholesterol, HDL-1 [mg/dL] | 8.2 |
| H2CH | Cholesterol, HDL-2 [mg/dL] | 9.2 |
| H3CH | Cholesterol, HDL-3 [mg/dL] | 6.1 |
| H4CH | Cholesterol, HDL-4 [mg/dL] | 4.7 |
| H1FC | Free Cholesterol, HDL-1 [mg/dL] | 13.4 |
| H2FC | Free Cholesterol, HDL-2 [mg/dL] | 18.3 |
| H3FC | Free Cholesterol, HDL-3 [mg/dL] | 13.4 |
| H4FC | Free Cholesterol, HDL-4 [mg/dL] | 7.3 |
| H1PL | Phospholipids, HDL-1 [mg/dL] | 12.2 |
| H2PL | Phospholipids, HDL-2 [mg/dL] | 10.7 |
| H3PL | Phospholipids, HDL-3 [mg/dL] | 8.0 |
| H4PL | Phospholipids, HDL-4 [mg/dL] | 2.0 |
| H1A1 * | Apo-A1, HDL-1 [mg/dL] | 15.0 |
| H2A1 | Apo-A1, HDL-2 [mg/dL] | 5.9 |
| H3A1 | Apo-A1, HDL-3 [mg/dL] | 5.3 |
| H4A1 | Apo-A1, HDL-4 [mg/dL] | 3.0 |
| H1A2 | Apo-A2, HDL-1 [mg/dL] | 23.6 |
| H2A2 | Apo-A2, HDL-2 [mg/dL] | 19.5 |
| H3A2 | Apo-A2, HDL-3 [mg/dL] | 12.0 |
| H4A2 | Apo-A2, HDL-4 [mg/dL] | 3.7 |

* Variables which have been excluded from analyses based on the spiking experiment.

Density ranges for lipoprotein main fractions: VLDL: 0.950-1.006 kg/L; IDL: 1.006-1.019 kg/L; LDL: 1.019-1.063 kg/L; HDL: 1.063-1.210 kg/L. Density ranges for lipoprotein subfractions: LDL1: 1.019-1.031 kg/L; LDL2: 1.031-1.034 kg/L; LDL3: 1.034-1.037 kg/L; LDL4: 1.037-1.040 kg/L; LDL5: 1.040-1.044 kg/L; LDL6: 1.044-1.063 kg/L; HDL1: 1.063-1.100 kg/L; HDL2: 1.100-1.112 kg/L; HDL3: 1.112-1.125 kg/L; HDL4: 1.125-1.210 kg/L.. TP: total plasma; VLDL: very-low density lipoprotein; IDL: intermediate-density lipoprotein; LDL: low-density lipoprotein; HDL: high-density lipoprotein; CH: cholesterol; FC: free cholesterol; PL: phospholipids; TG: triglycerides. AB: apolipoprotein-B; A1: apolipoprotein-1; A2: apolipoprotein-2.

**Table S3.** Concentrations (in mg/dL) of the different lipoprotein subfractions for the cases and controls in the study cohort. Values are given as mean (SD). SD: standard deviation.

| | | Premenopausal women | | Postmenopausal women | |
|---|---|---|---|---|---|
| | Lipoprotein subfraction | Case | Control | Case | Control |
| Total plasma | TPA1 * | 142.3 (19.3) | 142.8 (20.5) | 146.0 (19.6) | 145.2 (20.2) |
| | TPA2 | 29.4 (4.4) | 28.9 (4.0) | 29.5 (4.2) | 29.3 (4.6) |
| | TPAB * | 84.5 (18.7) | 86.5 (21.6) | 106.7 (22.7) | 107.4 (24.5) |
| | TPCH * | 207.3 (36.9) | 209.8 (40.4) | 245.6 (44.0) | 247.6 (46.6) |
| | TPTG * | 118.9 (59.9) | 128.4 (61.8) | 165.7 (76.3) | 167.7 (89.8) |
| VLDL | VLAB * | 7.8 (3.4) | 8.5 (3.6) | 10.8 (4.3) | 11.0 (4.9) |
| | VLCH * | 22.4 (11.4) | 24.3 (12.2) | 32.5 (14.7) | 33.6 (16.5) |
| | VLFC * | 9.8 (4.3) | 10.6 (4.5) | 13.5 (5.4) | 13.9 (6.1) |
| | VLPL * | 18.8 (8.3) | 20.4 (8.3) | 24.8 (10.2) | 25.3 (11.3) |
| | VLTG * | 63.5 (40.0) | 69.8 (40.8) | 91.2 (50.9) | 92.1 (59.7) |
| VLDL-1 | V1CH * | 7.7 (5.9) | 8.5 (6.1) | 11.7 (7.3) | 12.1 (8.8) |
| | V1FC * | 1.5 (1.8) | 1.7 (1.8) | 2.5 (2.2) | 2.6 (2.6) |
| | V1PL * | 4.4 (3.9) | 4.9 (4.0) | 6.6 (4.8) | 6.7 (5.5) |
| | V1TG * | 29.6 (27.3) | 32.9 (27.8) | 44.8 (34.0) | 45.0 (41.5) |
| VLDL-2 | V2CH * | 3.5 (1.9) | 3.8 (2.1) | 5.2 (2.5) | 5.3 (2.8) |
| | V2FC * | 1.5 (1.0) | 1.7 (1.1) | 2.3 (1.3) | 2.4 (1.5) |
| | V2PL * | 2.8 (1.5) | 3.0 (1.6) | 3.8 (1.9) | 3.9 (2.0) |
| | V2TG * | 12.2 (5.9) | 13.2 (6.2) | 16.8 (7.6) | 16.8 (7.9) |
| VLDL-3 | V3CH * | 3.9 (2.3) | 4.2 (2.5) | 5.7 (3.0) | 6.0 (3.3) |
| | V3FC * | 1.6 (1.0) | 1.8 (1.1) | 2.4 (1.4) | 2.5 (1.6) |
| | V3PL * | 3.3 (1.7) | 3.5 (1.9) | 4.5 (2.3) | 4.6 (2.5) |
| | V3TG * | 11.6 (5.3) | 12.5 (5.8) | 15.8 (7.1) | 16.1 (7.6) |
| VLDL-4 | V4CH * | 5.1 (2.3) | 5.4 (2.7) | 7.4 (3.2) | 7.8 (3.4) |
| | V4FC * | 2.3 (1.2) | 2.5 (1.4) | 3.5 (1.6) | 3.7 (1.8) |
| | V4PL * | 4.3 (1.7) | 4.6 (1.9) | 6.0 (2.2) | 6.2 (2.4) |
| | V4TG * | 8.0 (3.1) | 8.6 (3.4) | 10.7 (4.1) | 11.0 (4.4) |
| VLDL-5 | V5CH * | 1.2 (0.6) | 1.2 (0.6) | 1.3 (0.6) | 1.4 (0.7) |
| | V5FC * | 1.0 (0.6) | 1.1 (0.6) | 1.3 (0.7) | 1.4 (0.8) |
| | V5PL * | 1.9 (0.7) | 2.0 (0.7) | 2.2 (0.7) | 2.2 (0.7) |
| | V5TG * | 3.3 (0.8) | 3.3 (0.8) | 3.8 (0.8) | 3.8 (0.9) |
| IDL | IDAB * | 5.0 (2.5) | 5.2 (2.7) | 7.6 (3.2) | 7.8 (3.8) |
| | IDCH * | 13.5 (7.2) | 14.2 (8.0) | 21.3 (9.7) | 22.0 (11.1) |
| | IDFC * | 3.5 (2.0) | 3.7 (2.3) | 5.6 (2.7) | 5.8 (3.1) |
| | IDPL * | 5.6 (3.2) | 6.0 (3.4) | 8.6 (4.1) | 8.7 (4.5) |
| | IDTG * | 8.3 (8.7) | 9.5 (8.7) | 14.3 (11.0) | 14.2 (12.4) |
| LDL | LDAB * | 69.5 (15.3) | 70.5 (17.3) | 84.9 (18.0) | 84.9 (19.4) |
| | LDCH * | 111.2 (27.0) | 111.2 (27.7) | 131.2 (30.3) | 130.8 (32.1) |
| | LDFC * | 37.7 (7.7) | 37.6 (7.8) | 43.4 (8.5) | 43.3 (8.9) |
| | LDPL * | 62.9 (13.2) | 63.0 (13.9) | 73.0 (14.8) | 72.9 (16.0) |
| | LDTG | 18.7 (6.0) | 19.6 (7.3) | 25.6 (7.9) | 25.9 (9.0) |
| LDL-1 | L1AB | 12.8 (4.1) | 13.0 (4.5) | 16.4 (5.0) | 16.8 (5.2) |

| | | | | | |
|---|---|---|---|---|---|
| | L1CH * | 24.0 (8.2) | 24.0 (8.7) | 30.3 (9.9) | 31.1 (10.3) |
| | L1FC * | 7.5 (2.4) | 7.5 (2.6) | 9.4 (3.0) | 9.7 (3.1) |
| | L1PL * | 13.7 (4.2) | 13.7 (4.5) | 17.1 (5.1) | 17.5 (5.3) |
| | L1TG * | 5.7 (2.5) | 6.0 (3.0) | 8.4 (3.4) | 8.6 (3.8) |
| LDL-2 | L2TG * | 2.4 (0.8) | 2.4 (1.0) | 3.1 (1.0) | 3.1 (1.0) |
| LDL-3 | L3AB * | 12.0 (3.5) | 11.8 (3.5) | 13.4 (3.8) | 13.2 (4.1) |
| | L3CH * | 20.6 (6.6) | 20.0 (6.4) | 22.5 (7.1) | 22.1 (7.7) |
| | L3FC * | 6.9 (1.7) | 6.7 (1.6) | 7.4 (1.8) | 7.4 (2.0) |
| | L3PL * | 11.5 (3.3) | 11.2 (3.2) | 12.5 (3.6) | 12.3 (3.9) |
| | L3TG * | 2.7 (0.6) | 2.7 (0.8) | 3.3 (0.7) | 3.3 (0.7) |
| LDL-5 | L5AB * | 9.4 (3.5) | 9.9 (3.8) | 11.9 (4.4) | 11.8 (4.8) |
| | L5CH * | 13.5 (5.3) | 14.1 (5.5) | 16.6 (6.5) | 16.3 (7.0) |
| | L5FC * | 4.6 (1.4) | 4.7 (1.4) | 5.3 (1.6) | 5.3 (1.8) |
| | L5PL * | 7.7 (2.7) | 8.0 (2.8) | 9.2 (3.3) | 9.0 (3.6) |
| LDL-6 | L6AB * | 13.4 (5.4) | 14.2 (6.4) | 18.3 (7.4) | 18.6 (8.7) |
| | L6CH * | 15.1 (6.4) | 15.9 (7.4) | 20.1 (8.6) | 20.5 (9.6) |
| | L6FC * | 4.4 (1.6) | 4.4 (1.7) | 5.2 (1.8) | 5.2 (2.0) |
| | L6PL * | 9.2 (3.2) | 9.6 (3.7) | 11.5 (4.3) | 11.7 (4.8) |
| | L6TG * | 3.8 (1.3) | 3.9 (1.5) | 5.0 (1.7) | 5.1 (2.0) |
| HDL | HDA1 | 142.8 (20.7) | 143.2 (22.1) | 145.1 (21.4) | 144.1 (22.1) |
| | HDA2 * | 29.3 (4.2) | 28.8 (3.7) | 29.5 (3.9) | 29.4 (4.4) |
| | HDCH | 55.1 (10.9) | 54.7 (11.5) | 54.4 (12.1) | 54.3 (12.4) |
| | HDFC * | 18.5 (3.0) | 18.5 (3.3) | 19.7 (3.4) | 19.6 (3.5) |
| | HDPL | 76.0 (14.4) | 75.8 (15.1) | 75.5 (15.2) | 74.8 (15.5) |
| | HDTG * | 10.2 (4.1) | 10.8 (4.2) | 12.6 (4.0) | 12.6 (4.1) |
| HDL-1 | H1A2 | 2.8 (1.5) | 2.9 (1.6) | 3.0 (1.5) | 2.9 (1.5) |
| | H1CH | 17.8 (7.9) | 17.9 (8.7) | 18.5 (8.6) | 18.5 (8.6) |
| | H1FC * | 6.3 (1.8) | 6.3 (2.0) | 6.6 (2.0) | 6.6 (2.0) |
| | H1PL | 22.4 (9.8) | 22.7 (10.8) | 23.0 (10.4) | 22.8 (10.3) |
| | H1TG * | 3.2 (2.0) | 3.5 (2.3) | 4.3 (2.1) | 4.3 (2.2) |
| HDL-2 | H2A1 | 17.7 (4.4) | 17.8 (4.4) | 18.0 (4.4) | 17.7 (4.4) |
| | H2A2 * | 3.2 (1.2) | 3.2 (1.1) | 3.3 (1.1) | 3.3 (1.3) |
| | H2CH | 8.6 (2.3) | 8.5 (2.2) | 8.5 (2.3) | 8.5 (2.4) |
| | H2FC * | 2.6 (0.6) | 2.5 (0.6) | 2.7 (0.6) | 2.6 (0.6) |
| | H2PL | 13.2 (3.6) | 13.1 (3.4) | 13.2 (3.4) | 13.0 (3.5) |
| | H2TG * | 1.7 (0.8) | 1.8 (0.8) | 2.1 (0.8) | 2.1 (0.8) |
| HDL-3 | H3A1 * | 27.1 (4.6) | 27.0 (4.1) | 27.8 (4.4) | 27.7 (4.5) |
| | H3A2 * | 6.2 (1.4) | 6.1 (1.2) | 6.3 (1.3) | 6.3 (1.4) |
| | H3CH | 10.1 (1.7) | 9.8 (1.7) | 9.8 (1.8) | 9.8 (1.8) |
| | H3FC | 2.8 (0.6) | 2.7 (0.6) | 2.8 (0.6) | 2.8 (0.7) |
| | H3PL * | 15.3 (3.0) | 15.1 (2.8) | 14.8 (3.0) | 14.7 (3.0) |
| | H3TG * | 2.0 (0.8) | 2.0 (0.8) | 2.3 (0.8) | 2.3 (0.8) |
| HDL-4 | H4A1 | 69.1 (10.7) | 69.0 (10.1) | 69.8 (10.9) | 69.6 (12.3) |
| | H4A2 | 16.7 (4.0) | 16.3 (3.4) | 16.4 (3.4) | 16.4 (3.7) |
| | H4CH * | 17.5 (4.2) | 17.1 (3.9) | 16.5 (4.1) | 16.5 (4.6) |
| | H4FC | 4.5 (1.3) | 4.4 (1.1) | 4.5 (1.2) | 4.5 (1.3) |

| | | | | | |
|---|---|---|---|---|---|
| | H4PL * | 25.0 (4.9) | 24.7 (4.6) | 23.8 (4.7) | 23.6 (5.6) |
| | H4TG * | 3.4 (0.9) | 3.5 (0.9) | 3.9 (1.0) | 3.9 (1.0) |

TP: total plasma; VLDL: very-low density lipoprotein; IDL: intermediate-density lipoprotein; LDL: low-density lipoprotein; HDL: high-density lipoprotein; CH: cholesterol; FC: free cholesterol; PL: phospholipids; TG: triglycerides; AB: apolipoprotein-B; A1: apolipoprotein-1; A2: apolipoprotein-2.

* Significantly different between pre- and postmenopausal women.

**Table S4.** Sensitivity analyses on the found association between lipoprotein subfractions and overall breast cancer risk in premenopausal women of the HUNT2 study.

| | | Excluding women with reported use of HRT at HUNT2 or for whom this information was missing | Including only ER+ breast cases |
|---|---|---|---|
| | | OR (95% CI) | OR (95% CI) |
| Total plasma | TPA2 | 1.17 (0.97,1.41) | 1.14 (0.96,1.36) |
| | TPTG | 0.82 (0.66,1.02) | 0.78 (0.64,0.95) |
| VLDL | VLAB | 0.76 (0.62,0.94) | 0.77 (0.63,0.92) |
| | VLCH | 0.77 (0.62,0.95) | 0.79 (0.65,0.95) |
| | VLFC | 0.75 (0.61,0.93) | 0.78 (0.64,0.94) |
| | VLPL | 0.76 (0.62,0.92) | 0.79 (0.65,0.95) |
| | VLTG | 0.82 (0.66,1.02) | 0.79 (0.65,0.96) |
| VLDL-1 | V1CH | 0.88 (0.71,1.10) | 0.84 (0.69,1.01) |
| | V1FC | 0.90 (0.73,1.10) | 0.83 (0.69,0.99) |
| | V1PL | 0.91 (0.74,1.12) | 0.85 (0.71,1.02) |
| | V1TG | 0.92 (0.75,1.14) | 0.86 (0.71,1.03) |
| VLDL-2 | V2CH | 0.75 (0.61,0.93) | 0.76 (0.62,0.92) |
| | V2FC | 0.80 (0.65,0.99) | 0.78 (0.64,0.94) |
| | V2PL | 0.77 (0.63,0.94) | 0.76 (0.63,0.92) |
| | V2TG | 0.76 (0.62,0.93) | 0.75 (0.62,0.91) |
| VLDL-3 | V3CH | 0.77 (0.62,0.95) | 0.78 (0.64,0.94) |
| | V3FC | 0.78 (0.63,0.97) | 0.78 (0.64,0.95) |
| | V3PL | 0.75 (0.61,0.92) | 0.76 (0.62,0.92) |
| | V3TG | 0.72 (0.59,0.89) | 0.75 (0.61,0.91) |
| VLDL-4 | V4CH | 0.74 (0.60,0.90) | 0.81 (0.66,0.98) |
| | V4FC | 0.80 (0.64,0.98) | 0.81 (0.66,0.98) |
| | V4PL | 0.73 (0.60,0.90) | 0.77 (0.63,0.94) |
| | V4TG | 0.71 (0.58,0.87) | 0.76 (0.62,0.92) |
| VLDL-5 | V5CH | 0.75 (0.62,0.90) | 0.91 (0.77,1.08) |
| | V5PL | 0.75 (0.62,0.89) | 0.87 (0.73,1.03) |
| IDL | IDPL | 0.82 (0.66,1.03) | 0.81 (0.66,0.99) |
| | IDTG | 0.84 (0.68,1.04) | 0.79 (0.65,0.96) |
| LDL | LDTG | 0.86 (0.70,1.05) | 0.74 (0.60,0.91) |
| LDL-5 | L5AB | 0.80 (0.66,0.97) | 0.83 (0.69,1.00) |
| | L5CH | 0.81 (0.67,0.98) | 0.87 (0.72,1.04) |
| | L5PL | 0.81 (0.66,0.98) | 0.85 (0.71,1.02) |
| LDL-6 | L6AB | 0.78 (0.63,0.97) | 0.84 (0.69,1.03) |
| | L6TG | 0.80 (0.65,0.99) | 0.81 (0.66,0.99) |
| HDL | HDTG | 0.88 (0.74,1.04) | 0.79 (0.67,0.93) |
| HDL-1 | H1TG | 0.89 (0.76,1.05) | 0.83 (0.71,0.96) |
| HDL-2 | H2TG | 0.93 (0.79,1.09) | 0.81 (0.69,0.95) |
| HDL-3 | H3CH | 1.14 (0.97,1.35) | 1.12 (0.95,1.32) |
| | H3FC | 1.05 (0.88,1.26) | 1.09 (0.93,1.29) |
| HDL-4 | H4TG | 0.82 (0.69,0.98) | 0.83 (0.70,0.97) |

Results given as odds ratios and 95% confidence intervals (CI) per 1 SD increase for lipoproteins subfraction (1) excluding women with reported use of HRT at HUNT2 or for whom this information was missing and (2) including only ER+ breast cases and their matched controls. TP: total plasma; VLDL: very-low density lipoprotein; IDL: intermediate-density lipoprotein; LDL: low-density lipoprotein; HDL: high-density lipoprotein; CH: cholesterol; FC: free cholesterol; PL: phospholipids; TG: triglycerides; AB: apolipoprotein-B; A2: apolipoprotein-2.

**Table S5.** Odds ratios and 95% confidence intervals (CI) per 1 SD increase for metabolites significantly associated with risk of overall breast cancer (p-value < 0.05 and q-value < 0.05) in premenopausal women of the HUNT2 study.

| | Baseline model* | | | Adjusted model** | | |
|---|---|---|---|---|---|---|
| Metabolite | OR (95% CI) | P | $P_{adj}$ | OR (95% CI) | P | $P_{adj}$ |
| Formate | 0.99 (0.86,1.14) | 0.89 | 1.00 | 1.00 (0.87,1.14) | 0.96 | 0.97 |
| Creatine | 1.00 (0.87,1.15) | 0.99 | 1.00 | 1.01 (0.88,1.17) | 0.84 | 0.97 |
| Lactate | 1.00 (0.87,1.15) | 1.00 | 1.00 | 1.01 (0.87,1.17) | 0.88 | 0.97 |
| Glycine | 1.00 (0.87,1.14) | 0.95 | 1.00 | 1.00 (0.88,1.15) | 0.95 | 0.97 |
| Methanol | 0.99 (0.87,1.13) | 0.94 | 1.00 | 1.00 (0.88,1.14) | 0.96 | 0.97 |
| Dimethyl-sulfone | 1.06 (0.90,1.23) | 0.49 | 1.00 | 1.08 (0.92,1.26) | 0.36 | 0.97 |
| Ornithine | 1.00 (0.87,1.15) | 1.00 | 1.00 | 1.00 (0.86,1.15) | 0.97 | 0.97 |
| Methionine | 1.00 (0.86,1.17) | 0.96 | 1.00 | 0.99 (0.85,1.16) | 0.90 | 0.97 |
| Glutamine | 0.96 (0.84,1.10) | 0.60 | 1.00 | 0.95 (0.83,1.10) | 0.50 | 0.97 |
| Citrate | 0.99 (0.84,1.16) | 0.87 | 1.00 | 1.03 (0.88,1.21) | 0.71 | 0.97 |
| **Acetate** | **1.38 (1.04,1.89)** | **0.04** | **0.67** | **1.42 (1.05,1.95)** | **0.03** | **0.45** |
| Acetoacetate | 1.03 (0.89,1.20) | 0.67 | 1.00 | 1.03 (0.89,1.21) | 0.68 | 0.97 |
| Glutamate | 1.00 (0.86,1.17) | 0.98 | 1.00 | 1.03 (0.87,1.22) | 0.72 | 0.97 |
| Pyruvate | 0.90 (0.79,1.02) | 0.10 | 0.67 | 0.90 (0.79,1.02) | 0.10 | 0.85 |
| Alanine | 0.88 (0.77,1.01) | 0.08 | 0.67 | 0.89 (0.78,1.03) | 0.12 | 0.85 |
| Ethanol | 1.05 (0.94,1.22) | 0.40 | 1.00 | 1.06 (0.95,1.23) | 0.34 | 0.97 |
| Isoleucine | 0.91 (0.78,1.06) | 0.22 | 1.00 | 0.92 (0.78,1.08) | 0.32 | 0.97 |
| 2-Methylglutarate | 0.99 (0.84,1.16) | 0.88 | 1.00 | 0.98 (0.83,1.16) | 0.82 | 0.97 |
| Leucine | 0.92 (0.78,1.08) | 0.32 | 1.00 | 0.94 (0.79,1.12) | 0.49 | 0.97 |
| **Phenylalanine** | 1.14 (0.97,1.34) | 0.11 | 0.67 | **1.20 (1.02,1.43)** | **0.03** | **0.45** |
| Glucose | 1.10 (0.91,1.34) | 0.33 | 1.00 | 1.11 (0.91,1.36) | 0.29 | 0.97 |
| Tyrosine | 0.98 (0.85,1.13) | 0.78 | 1.00 | 0.98 (0.85,1.14) | 0.82 | 0.97 |
| Creatinine | 0.96 (0.81,1.13) | 0.60 | 1.00 | 0.97 (0.82,1.16) | 0.76 | 0.97 |
| Valine | 0.99 (0.84,1.15) | 0.86 | 1.00 | 1.02 (0.87,1.20) | 0.81 | 0.97 |
| Proline-betaine | 1.06 (0.93,1.20) | 0.39 | 1.00 | 1.07 (0.94,1.21) | 0.29 | 0.97 |
| Histidine | 0.98 (0.85,1.13) | 0.79 | 1.00 | 0.99 (0.86,1.15) | 0.92 | 0.97 |
| Lysine | 0.99 (0.86,1.15) | 0.94 | 1.00 | 1.02 (0.87,1.19) | 0.80 | 0.97 |
| 3-Hydroxybutyrate | 1.13 (0.97,1.33) | 0.12 | 0.67 | 1.12 (0.95,1.32) | 0.18 | 0.97 |

*Baseline model: adjusted for lab for NMR analyses and matching variable (Age at participation in the HUNT2 study)

**Adjusted model: in addition to lab and age, this model is adjusted for no. of full-term pregnancies, age at menarche, BMI, alcohol consumption and smoking status. Bold font indicates variables significant in the adjusted model after multiple testing correction.

#### Supplementary Figures

**Figure S1.** Results from the spiking experiment for donor 3 (top) and donor 6 (bottom). The green lines show the spectra from the clean serum samples from donors 3 and 6, the red lines show the spectra of the serum samples from the donors spiked with 0.2 mM neopentyl glycol, and the blue lines show the processed spiked spectra, where the signal from neopentyl glycol has been removed.

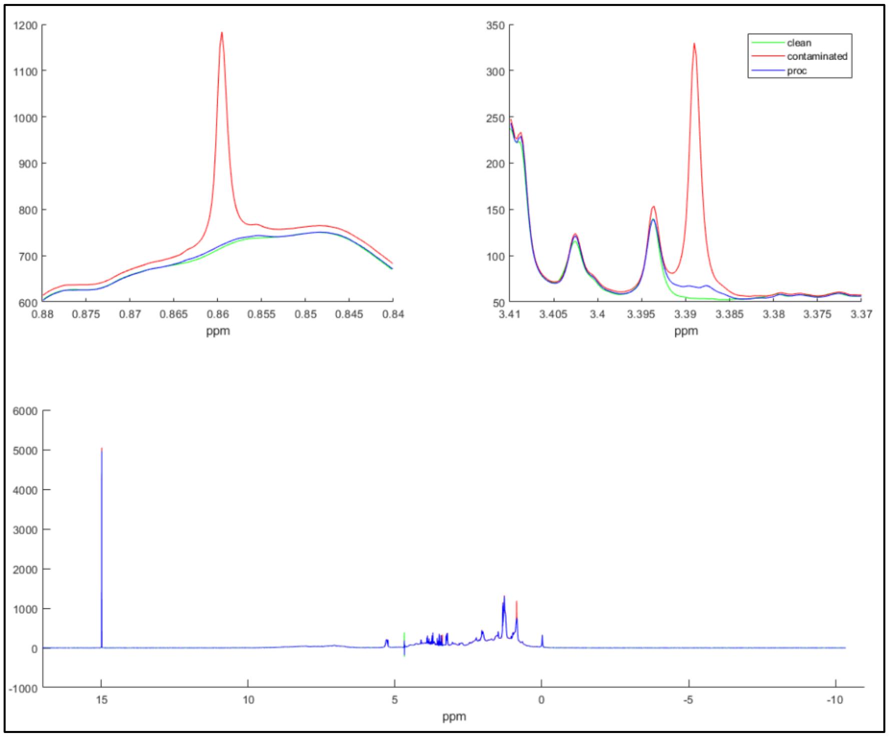


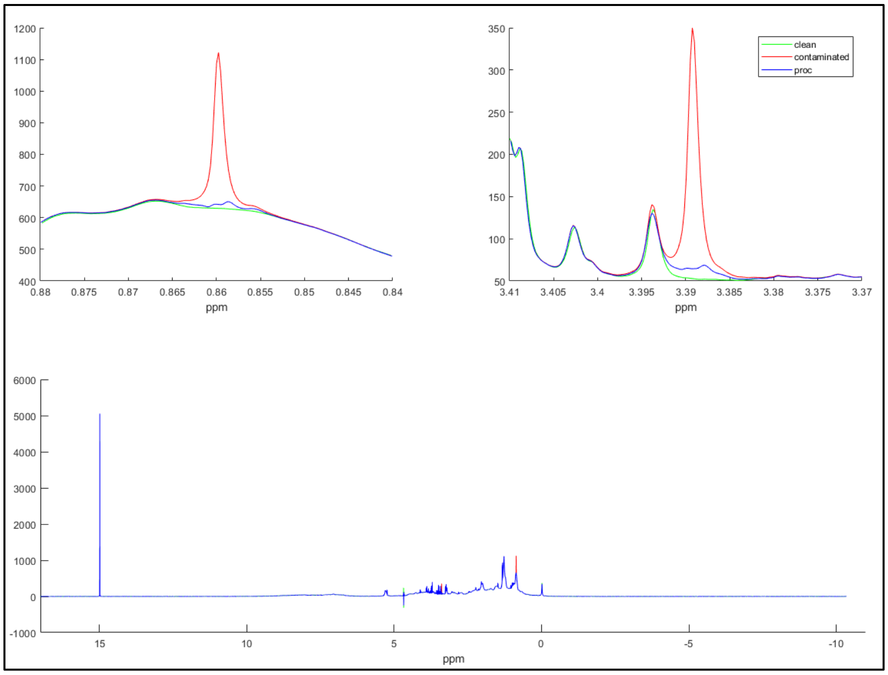

**Figure S2.** Correlation matrix of the metabolites and lipoprotein subfractions included in this study. The color of the circles shows the direction of the correlation (blue positive and red negative), while the size of the circles shows the size of the correlation.

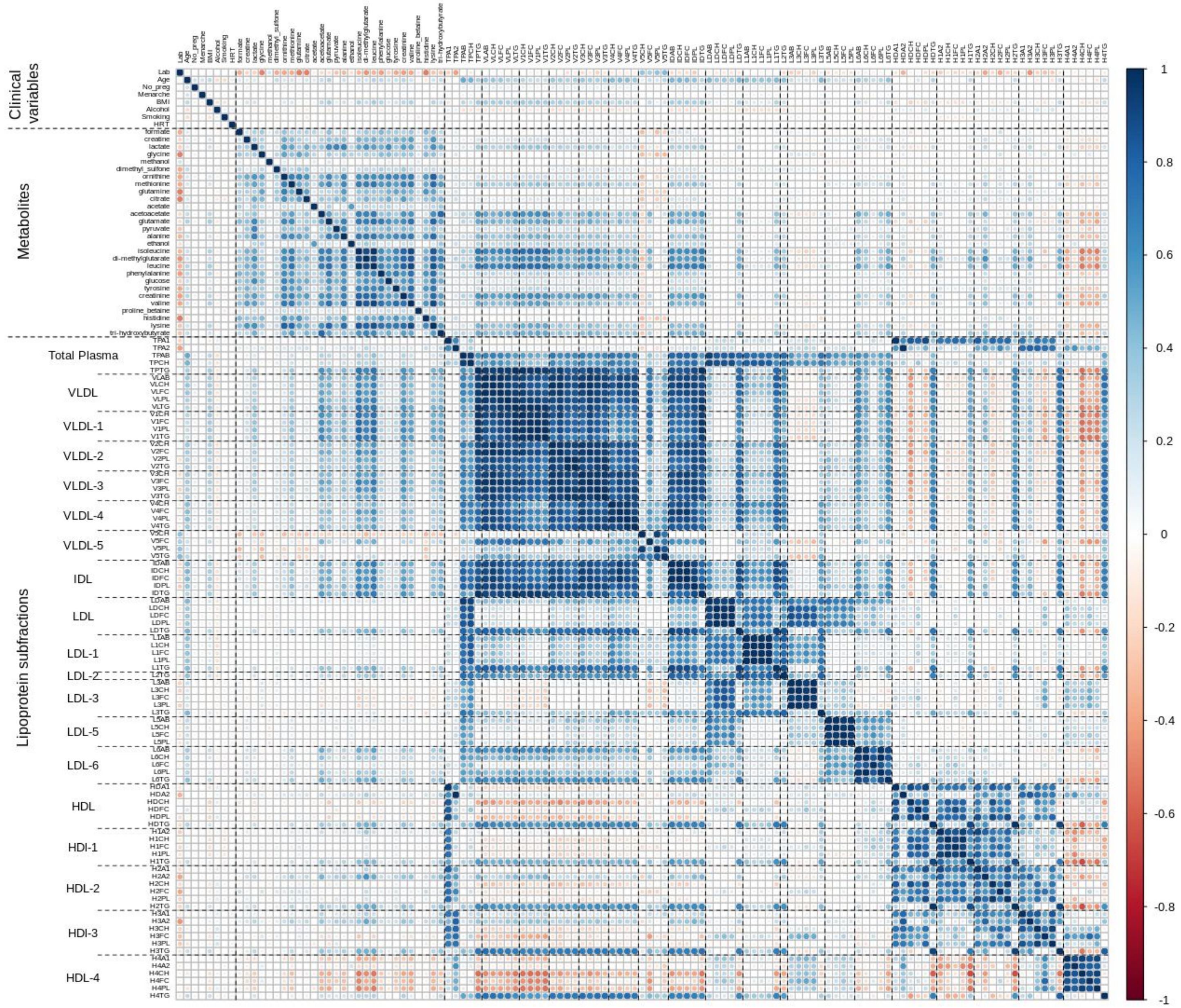
